# On the relationship between serial interval, infectiousness profile and generation time

**DOI:** 10.1101/2020.09.18.20197210

**Authors:** Sonja Lehtinen, Peter Ashcroft, Sebastian Bonhoeffer

## Abstract

The timing of transmission plays a key role in the dynamics and controllability of an epidemic. However, observing the distribution of generation times (time interval between the points of infection of an infector and infectee in a transmission pair) requires data on infection times, which are generally unknown. The timing of symptom onset is more easily observed; the generation time distribution is therefore often estimated based on the serial interval distribution (distribution of time intervals between symptom onset of an infector and an infectee). This estimation follows one of two approaches: i) approximating the generation time distribution by the serial interval distribution; or ii) deriving the generation time distribution from the serial interval and incubation period (time interval between infection and symptom onset in a single individual) distributions. These two approaches make different – and not always explicitly stated – assumptions about the relationship between infectiousness and symptoms, resulting in different generation time distributions with the same mean but unequal variances. Here, we clarify the assumptions that each approach makes and show that neither set of assumptions is plausible for most pathogens. However, the variances of the generation time distribution derived under each assumption can reasonably be considered as upper (approximation with serial interval) and lower (derivation from serial interval) bounds. Thus, we suggest a pragmatic solution is to use both approaches and treat these as edge cases in downstream analysis. We discuss the impact of the variance of the generation time distribution on the controllability of an epidemic through strategies based on contact tracing, and we show that underestimating this variance is likely to overestimate controllability.

## Background

### Motivation

Estimating the generation time (the timing between successive infections in a transmission chain) distribution in an emerging epidemic is both extremely important and extremely challenging. Generation time is key to assessing the controllability of the epidemic: it determines the relationship between the basic reproductive number *R*_0_ and the epidemic’s growth rate [1, 2], as well as how much delays in the isolation of infected individuals impede epidemic control [3, 4]. However, the timing of transmission events is often unknown. The distribution of generation times is therefore typically estimated based on the timing of symptom onset, which requires assumptions about the relationship between infectiousness and symptoms. These assumptions are not always explicitly stated and their plausibility is rarely discussed. Here, we illustrate how assumptions about infectiousness and symptom onset affect the relationship between the generation time and serial interval distributions, and the implications this has for assessing epidemic controllability.

### Definitions

We consider an infector *i* and infectee *j* (Figure 1A), and define: *S*_*ij*_ as the serial interval (time interval between symptom onset of infector *i* and symptom onset of infectee *j*); *G*_*ij*_ as the generation time (time interval from infection of *i* to infection of *j*); *P*_*ij*_ as the time interval from symptom onset of *i* to infection of *j*; and *I*_*i*_ as the incubation period of *i* (and *I*_*j*_ is the incubation period of *j*). We use the letters without indices to denote the distributions of these time intervals (e.g. *S* is the serial interval distribution). The generation time distribution *G* describes infectiousness relative to the point of infection, while *P* describes infectiousness relative to symptom onset. We refer to *P* as the infectiousness profile [5, 6].

**Figure 1:**
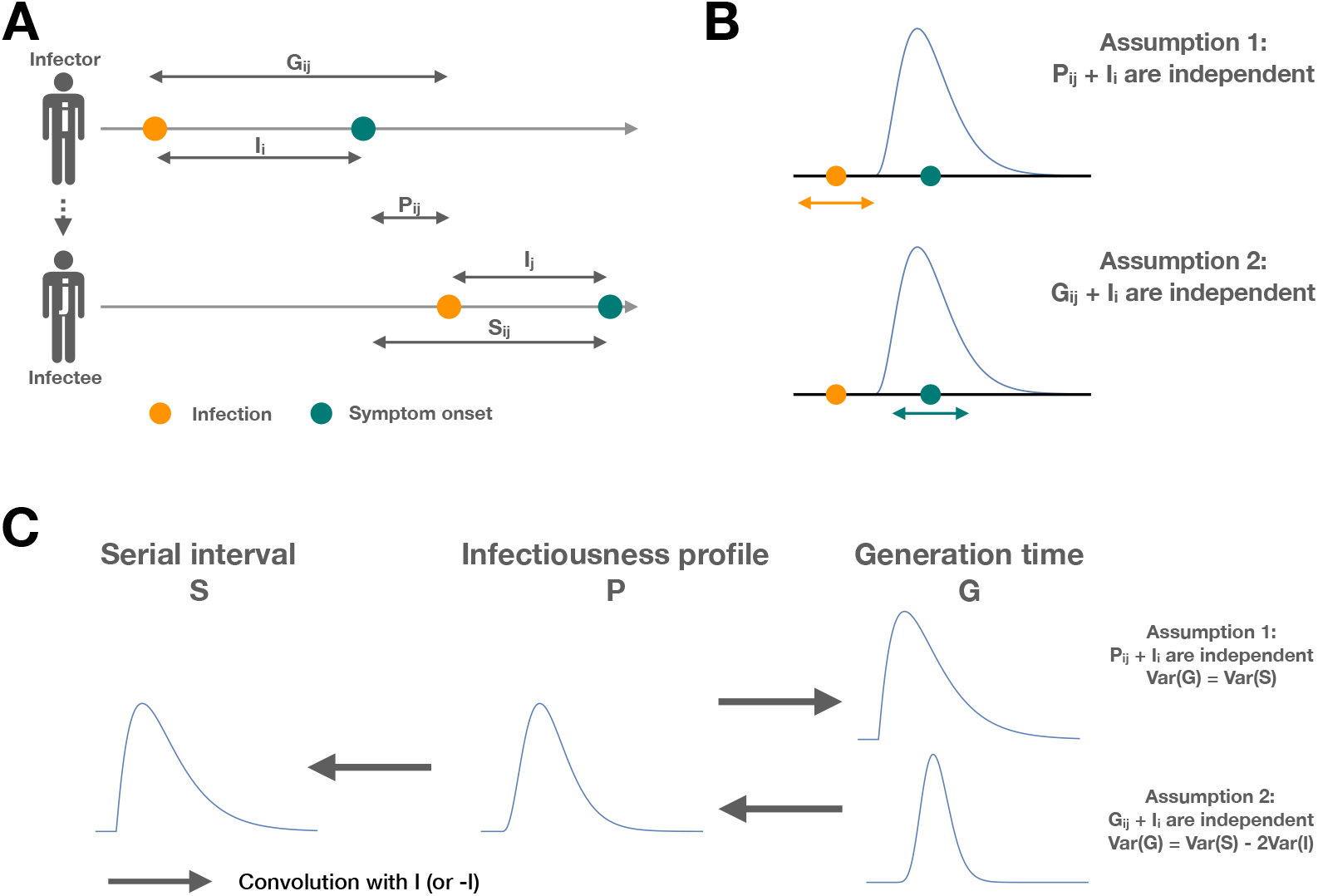
A schematic of how the assumptions about infectiousness and symptoms affect the relationship between the serial interval and generation time distributions. A: Definitions of: serial interval *S*_*ij*_, time from symptom onset of infector *i* to symptom onset of infectee *j*; generation time *G*_*ij*_, time from infection of *i* to infection of *j*; incubation time *I*_*i*_, time from infection of *i* to symptom onset of *i*; and *P*_*ij*_, time from symptom onset of *i* to infection of *j*. B: Illustration of how infectiousness relates to the point of infection and onset of symptoms under the two different assumptions. Under assumption 1 (*P*_*ij*_ and *I*_*i*_ independent), the infectiousness is fixed with reference to symptom onset. Under assumption 2 (*G*_*ij*_ and *I*_*i*_ independent), the infectiousness is fixed with reference to the point of infection. C: The relationship between the generation time distribution, the infectiousness profile, and the serial interval distribution under assumptions 1 and 2.

### Data availability in early epidemics

The distributions *I, G, P* and *S* are typically derived from contact tracing data during epidemic outbreaks. Such data consist of transmission pairs, usually with the timing of symptom onset for infector and infectee, and an exposure window for the timing of the transmission event. These data allow direct estimations *I* and *S*, but not *P* and *G*.

It is important to note that in a growing epidemic, there is a difference between distributions derived from backwards (from infectee to infector) and forwards (from infector to infectee) intervals. This is because in a growing epidemic, a randomly sampled infected is likely to have been infected more recently than expected based on the generation time. As contact tracing data typically gives rise to backward intervals, it is important to correct for this bias [2]. In this work, for clarity, we omit this correction when discussing the relationship between *G, P* and *S*.

### Relationship between *G, P* and *S*

#### Deriving *G* and *P* from *S*

The relationships between *G*_*ij*_, *P*_*ij*_ and *S*_*ij*_ are illustrated in Figure 1A and are captured by the following equations: 

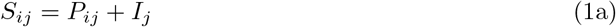

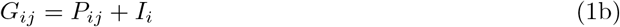

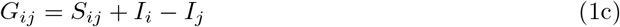

Deriving *P* from *S* does not require strong assumptions; *P*_*ij*_ and *I*_*j*_ are plausibly independent: it is reasonable to assume that the interval between the infector’s symptom onset and onward transmission does not affect the incubation period of the infectee. From Eq. (1a) we can then write *S* as the convolution between *P* and *I*, i.e. 

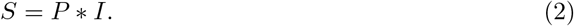

The infectiousness profile *P* can therefore be derived by deconvolution of the serial interval and incubation period distributions [5, 6].

Deriving *G* is not as straightforward: as *P*_*ij*_ and *I*_*i*_ relate to the same individual, independence of the two is a more debatable assumption than for *P*_*ij*_ and *I*_*j*_. Assuming *I*_*i*_ and *I*_*j*_ are independent and identically distributed (i.i.d), *S* and *G* have the same mean and their variances are related through [7]: 

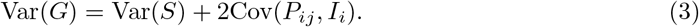

Deriving the generation time distribution *G* requires further assumptions: typically, either the independence of *P*_*ij*_ and *I*_*i*_ or the independence of *G*_*ij*_ and *I*_*i*_.

### Assumption 1: independence of incubation period of the infector (*I*_*i*_) and time from symptoms of infector to infection of infectee (*P*_*ij*_)

Under this assumption, there is no correlation between how long it takes an individual to develop symptoms and the interval between symptom onset and onward transmission (Figure 1B). Such a situation would arise, for example, if individuals have a variable period between infection and the onset of infectiousness, without this period affecting subsequent infectiousness or onset of symptoms (see also [2]).

Using Eq. (1b), the independence of *P*_*ij*_ and *I*_*i*_ means that *G* can be derived as the convolution of *P* and *I* (i.e. *G* = *P∗I*) and is thus identical to *S* (Eq. 2). Thus, the often-used approach of approximating the generation time distribution by the serial interval implicitly makes this assumption (Figure 1C).

In line with the above, under this assumption, the variance of *G* is equal to the variance of *S*: 

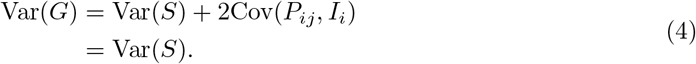

This assumption is biologically implausible: it requires the incubation period to be independent of processes affecting infectiousness. Yet infectiousness and symptom onset both likely depend on pathogen load; it is therefore unlikely that assumption 1 holds for most pathogens. Furthermore, unlike serial intervals, generation times cannot be negative. When observed, negative serial intervals are empirical evidence against assumption 1.

### Assumption 2: independence of incubation period of the infector (*I*_*i*_) and time from infection of infector to infection of infectee (*G*_*ij*_)

Under this assumption, the timing of transmission is uncorrelated with the timing of symptom onset (Figure 1B). As *P*_*ij*_ = *G*_*ij*_ − *I*_*i*_ [Eq. (1b)], *P* would then be the convolution of *G* and −*I, P* = *G ∗* (−*I*). The generation time distribution *G* could therefore be derived from *S* by deconvolving first with *I* and then with −*I* (Figure 1C), i.e. solving *S ∗I* = *G∗* (−*I*) for *G*. The functional form of *G* would therefore depend on both empirical distributions *S* and *I*. This is the approach adopted for deriving the generation interval of SARS-CoV-2 in Ferretti et al. [4] and Ganyani et al. [8].

In line with the above, under this assumption, the variance of *G* is smaller than the variance of *S*: 

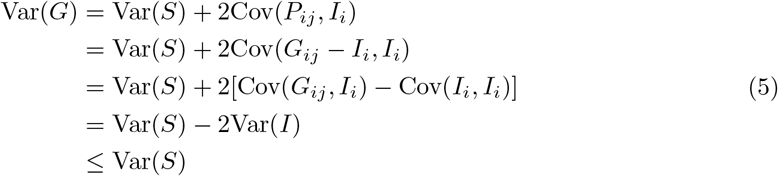

This assumption is also biologically implausible. If infectiousness and symptom onset both depend on pathogen load, individuals with a rapid increase in pathogen load will develop symptoms early (short *I*_*i*_) and transmit sooner after infection (small *G*_*ij*_), leading to Cov(*G*_*ij*_, *I*_*i*_) > 0. Furthermore, symptom onset itself is likely to affect infectiousness. Depending on the pathogen, the effect could be in either direction (symptomatic individuals transmitting more because symptoms contribute to transmission, or symptomatic individuals transmitting less because they self-isolate). However, either scenario would lead to a positive correlation between the timing of symptom onset and transmission (SI Figure 1).

### Assumptions 1 and 2 bound the variance of *G*

Although neither assumption 1 or 2 are plausible, they are still informative: the variances of the generation time distribution derived under these assumptions can reasonably be considered as upper and lower bounds for Var(*G*): 

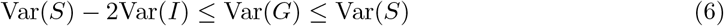

Assumption 1 leads to the upper bound Var(*G*) = Var(*S*). A greater variance would require Cov(*P*_*ij*_, *I*_*i*_) > 0 (see Eq. (3)): i.e. transmission occurring late with reference to symptoms for individuals with a longer incubation period - for example, a greater proportion of transmission being post-symptomatic when symptoms appear late. The notion that Cov(*P*_*ij*_, *I*_*i*_) > 0 is unlikely has also been previously suggested in the literature [2]. Furthermore, if negative serial intervals are observed, this suggests Var(*G*) < Var(*S*) (assuming the serial interval distribution and generation time distribution have a similar shape), since the distributions have the same mean and negative generation times are not possible.

Assumption 2 leads to the lower bound Var(*G*) = Var(*S*) − 2Var(*I*). A lower variance would require Cov(*G*_*ij*_, *I*_*i*_) < 0 (see Eq. (5)): i.e. transmission occurring soon after infection for individuals with a longer incubation period. However, as discussed above, individuals with a faster increase in pathogen load are likely to start transmitting earlier and also have shorter incubation period, leading to Cov(*G*_*ij*_, *I*_*i*_) > 0. Furthermore, if the appearance of symptoms leads to a change in infectiousness (in either direction), earlier symptoms will correlate with earlier transmission, again leading to Cov(*G*_*ij*_, *I*_*i*_) > 0.

### Possible solutions

#### Empirical testing of assumptions

*A priori*, there is no reason to consider either assumption 1 or assumption 2 as more plausible that the other. With appropriate data, the assumptions can be tested empirically. For example, such analysis for SARS-CoV-2 suggests a strong positive correlation between *G*_*ij*_ and *I*_*i*_, and a weak negative correlation between *P*_*ij*_ and *I*_*i*_ [9]. In other words, for SARS-CoV-2, neither assumption holds, but assumption 1 (independence of *P*_*ij*_ and *I*_*i*_) is a better approximation.

The empirical testing of these assumptions requires transmission pairs for which *I*_*i*_ and *G*_*ij*_ (or, equivalently, *P*_*ij*_) can be estimated. This can be done with either: i) data on the timing of infection for both infector *i* and infectee *j* and the timing of symptom onset for *i*; or ii) data on the timing of symptom onset for both *i* and *j* and the timing infection for *i*, as the assumption that *P*_*ij*_ and *I*_*j*_ are independent allows estimating the timing of infection for *j*. Therefore, an interesting corollary here is that for transmission pairs with a known serial interval, data on the timing of infection of the infector is more informative than the timing of infection of the infectee.

In practice, when such data are available, the generation interval distribution can simply be directly estimated from the data. The reason for deriving *G* from *S* is precisely the lack of such data; an alternative approach for assessing the plausibility of the assumptions underlying this derivation is therefore necessary.

### Assumptions 1 and 2 as edge cases

As assumptions 1 and 2 bound the variance of *G*, a solution when data are lacking is to derive *G* under both assumptions, and treat these as boundary cases in downstream analysis (e.g. best and worst case scenarios). This approach may not always be entirely straightforward. If Var(*S*) < 2Var(*I*), assumption 2 would lead to negative variance of *G*. In these cases, the lower bound for Var(*G*) is zero. If the serial interval distribution includes negative values, deriving *G* under assumption 1 is problematic. A pragmatic approach in these cases would be to use Var(*G*) = Var(*S*) and to assume a non-negative functional form for *G* (e.g. lognormal, gamma or Weibull), although the resulting distribution will not be the correct distribution under assumption 1. The key point is that evidence against assumption 1, such as negative serial intervals, is not, in itself, evidence in favour of assumption 2.

## Implications for the modelling of contact tracing

Finally, we explore the impact of the variance and functional form of the generation time distribution on the modelling of contact tracing, using the example of SARS-CoV-2. Table 1 shows empirical estimates for the mean and variance of the serial interval and incubation period distributions. Both distributions have a mean of around 5 days. The variance of the incubation period distribution is generally estimated to be in the range of 5 to 8 days^2^, although some studies have also reported considerably higher values (Table 1). With the exception of some smaller studies, the variance of the serial interval distribution is generally estimated to be of the order of 20 to 30 days^2^. Assuming Var(*S*) = 25 and Var(*I*) = 8, a plausible range for Var(*G*) would thus be 25 to 9 days^2^ under assumptions 1 and 2 respectively (though lower values cannot be excluded if the lower estimates of Var(*S*) hold).

**Table 1:** Shape, mean and variance of incubation period and serial interval distributions of SARS-CoV-2 from a range of studies. N indicates the sample size.

| Study | Distribution | Shape | Mean<br>(days) | Variance<br>(days<sup>2</sup>) | N |
| --- | --- | --- | --- | --- | --- |
| Zhang [10] | Incubation | Lognormal | 5.2 | 6.7 | 49 |
| Li [11] | Incubation | Lognormal | 5.2 | 15.3 | 10 |
| Lauer [12] | Incubation | Lognormal | 5.5 | 5.8 | 181 |
| Backer [13] | Incubation | Weibull | 6.4 | 5.3 | 88 |
| Linton [14] | Incubation | Lognormal | 5.6 | 7.8 | 158 |
| Ganyani [8] | Serial interval (Singapore) | Gamma | 5.2 | 18.7 | 54 |
| Ganyani [8] | Serial interval (Tianjin) | Gamma | 3.95 | 18.0 | 114 |
| Zhang [10] | Serial interval | Gamma | 5.1 | 7.2 | 34 |
| Li [11] | Serial interval | Gamma | 7.5 | 11.6 | 6 |
| He [5] | Serial interval | Gamma (shifted) | 5.8 | 20.0 | 77 |
| Nishiura [15] | Serial Interval | Lognormal | 4.7 | 8.4 | 28 |
| Ali [16] | Serial Interval (all) | Normal | 5.1 | 28.1 | 677 |
| Ali [16] | Serial Interval (pre-peak) | Normal | 7.1 | 27.0 | 162 |

Figure 2 illustrates how the variance and functional form of the generation time distribution impact how quickly infected individuals need to be isolated to prevent a significant portion of onward transmission, that is, how quickly contact tracing needs to operate for the epidemic to be controllable. For example, assuming generation time is gamma distributed with a mean of 5 days, preventing 80% of onward transmission requires isolation of an infected individual within 1.1 days if Var(*G*) is 25 days^2^, and 2.5 days if Var(*G*) is 9 days^2^. On the other hand, if the variance is large, isolating individuals even with considerable delay will still have an impact on onward transmission. For example, isolating an infected individual 10 days after infection will prevent 14% of onward transmission if Var(*G*) is 25 days^2^, but only 7% if Var(*G*) is 9 days^2^. In practice, if the goal of contact tracing is to control the epidemic, the former scenario is more relevant [4]. Thus underestimating the variance of the generation time distribution (assumption 2 risks overestimating the effectiveness of contact tracing.

**Figure 2:**
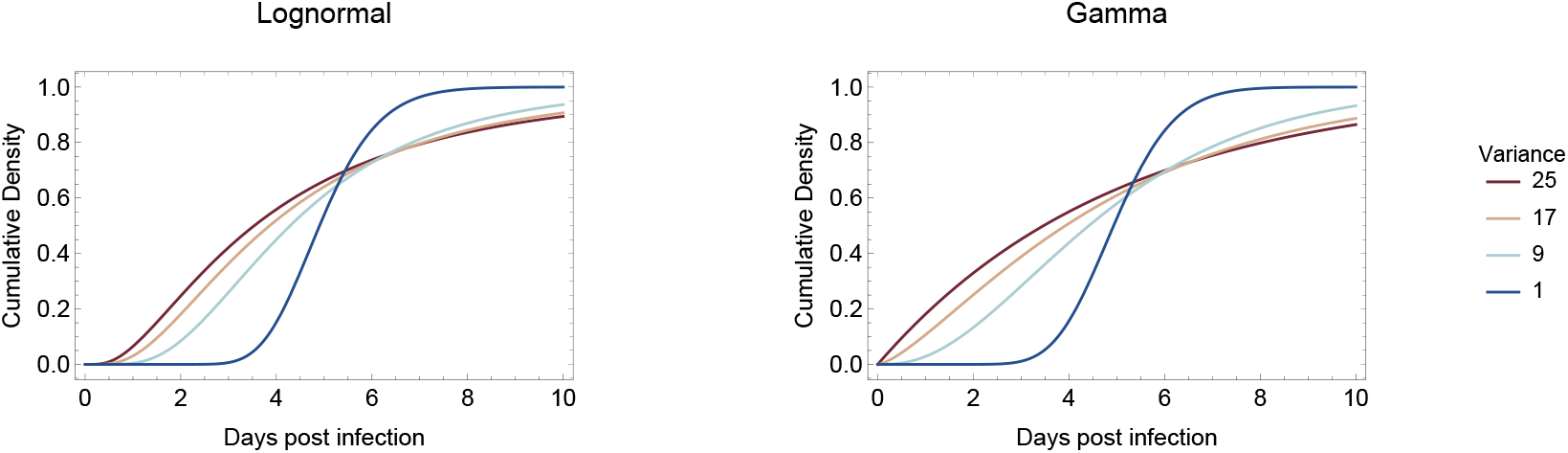
A schematic showing the impact of functional form and variance on the timing of onward transmission. The plots show cumulative generation time distributions, i.e. the proportion of transmission occurring within *x* days of infection. All distributions have a mean of 5 days.

## Conclusion

Neither of these two commonly used approaches for estimating the generation time distribution from the serial interval distribution is based on plausible assumptions for most pathogens. The two approaches yield generation time distributions with the same mean, but different variances. This difference in variance can have a considerable impact on estimating the controllability of an epidemic through contact tracing. The two variances are plausible upper and lower bounds for the variance of the generation time distribution. We therefore suggest a pragmatic solution is to treat the distributions derived through the two approaches as edge cases in downstream analysis. Preferring one approach over the other risks either over- or underestimating the controllability of an epidemic.

## Data Availability

All data generated or analysed during this study are included in this published article (and its supplementary information files).

## Acknowledgements

We thank Luca Ferretti and Jana Huisman for helpful discussion.

## Supporting Figure

**Figure 1:**
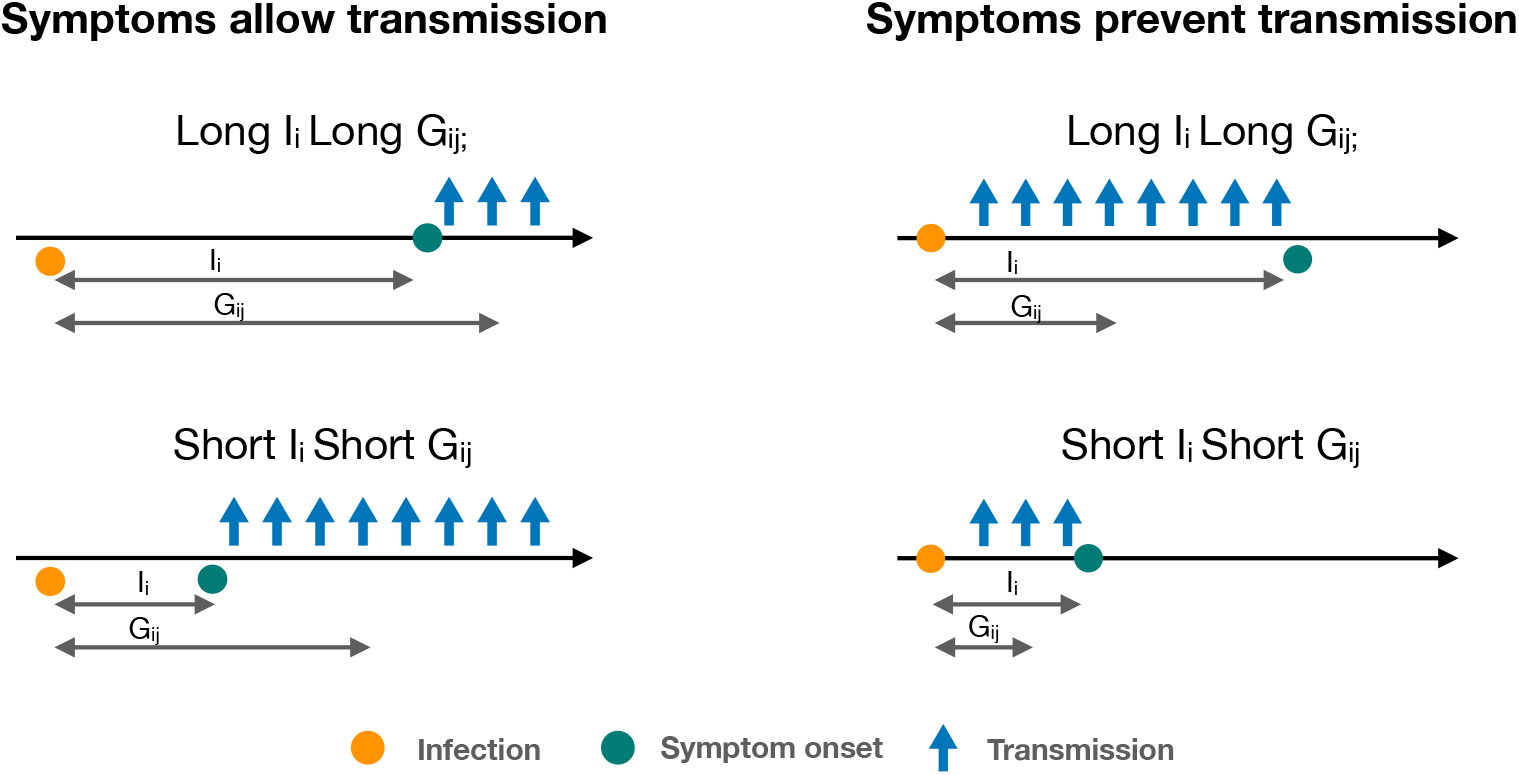
Schematic illustrating how symptom onset affecting transmission leads to a positive correlation between incubation period and generation time, whether symptoms increase or decrease transmission. In the left-hand panels, transmission only occurs after symptom onset. In the right-hand panels, transmission only occurs prior to symptom onset. We make the additional assumption that the length of the incubation period does not affect when infectiousness ends (for the left-hand panels) or starts (for the right-hand panel).

## Notes

### Competing Interest Statement

The authors have declared no competing interest.

### Funding Statement

This work was supported by the Swiss National Science Foundation. The funders of the study had no role in study design, data collection, data analysis, data interpretation, or writing of the manuscript.

### Author Declarations

No IRB needed

## References

[1] Wallinga J, Lipsitch M. How generation intervals shape the relationship between growth rates and reproductive numbers. Proceedings of the Royal Society B: Biological Sciences. 2007;274(1609):599–604.

[2] Britton T, Scalia Tomba G. Estimation in emerging epidemics: Biases and remedies. Journal of the Royal Society Interface. 2019;16(150):20180670.

[3] Fraser C, Riley S, Anderson RM, Ferguson NM. Factors that make an infectious disease outbreak controllable. Proceedings of the National Academy of Sciences. 2004;101(16):6146– 6151.

[4] Ferretti L, Wymant C, Kendall M, Zhao L, Nurtay A, Abeler-Dörner L, et al. Quantifying SARS-CoV-2 transmission suggests epidemic control with digital contact tracing. Science. 2020;368(6491).

[5] He X, Lau EH, Wu P, Deng X, Wang J, Hao X, et al. Temporal dynamics in viral shedding and transmissibility of COVID-19. Nature Medicine. 2020;26(5):672–675.

[6] Ashcroft P, Huisman JS, Lehtinen S, Bouman JA, Althaus CL, Regoes RR, et al. COVID-19 infectivity profile correction. Swiss Medical Weekly. 2020;150(3132).

[7] Svensson å. A note on generation times in epidemic models. Mathematical Biosciences. 2007;208(1):300–311.

[8] Ganyani T, Kremer C, Chen D, Torneri A, Faes C, Wallinga J, et al. Estimating the generation interval for coronavirus disease (COVID-19) based on symptom onset data, March 2020. Eurosurveillance. 2020;25(17):2000257.

[9] Ferretti L, Ledda A, Wymant C, Zhao L, Ledda V, Abeler-Dorner L, et al. The timing of COVID-19 transmission. medRxiv. 2020;Publisher: Cold Spring Harbor Laboratory Press eprint: https://www.medrxiv.org/content/early/2020/09/07/2020.09.04.20188516.full.pdf. xAvailable from: https://www.medrxiv.org/content/early/2020/09/07/2020.09.04.20188516.

[10] Zhang J, Litvinova M, Wang W, Wang Y, Deng X, Chen X, et al. Evolving epidemiology and transmission dynamics of coronavirus disease 2019 outside Hubei province, China: a descriptive and modelling study. The Lancet Infectious Diseases. 2020;.

[11] Li Q, Guan X, Wu P, Wang X, Zhou L, Tong Y, et al. Early transmission dynamics in Wuhan, China, of novel coronavirus–infected pneumonia. New England Journal of Medicine. 2020;.

[12] Lauer SA, Grantz KH, Bi Q, Jones FK, Zheng Q, Meredith HR, et al. The incubation period of coronavirus disease 2019 (COVID-19) from publicly reported confirmed cases: estimation and application. Annals of internal medicine. 2020;172(9):577–582.

[13] Backer JA, Klinkenberg D, Wallinga J. Incubation period of 2019 novel coronavirus (2019-nCoV) infections among travellers from Wuhan, China, 20–28 January 2020. Eurosurveillance. 2020;25(5):2000062.

[14] Linton NM, Kobayashi T, Yang Y, Hayashi K, Akhmetzhanov AR, Jung Sm, et al. Incubation period and other epidemiological characteristics of 2019 novel coronavirus infections with right truncation: a statistical analysis of publicly available case data. Journal of Clinical Medicine. 2020;9(2):538.

[15] Nishiura H, Linton NM, Akhmetzhanov AR. Serial interval of novel coronavirus (COVID-19) infections. International Journal of Infectious Diseases. 2020;.

[16] Ali ST, Wang L, Lau EH, Xu XK, D.Z, Wu Y, et al. Serial interval of SARS-CoV-2 was shortened over time by nonpharmaceutical interventions. Science. 2020;.

